# Prevalence and Predictors of General Psychiatric Disorders and Loneliness during COVID-19 in the United Kingdom: Results from the Understanding Society UKHLS

**DOI:** 10.1101/2020.06.09.20120139

**Authors:** Lambert Zixin Li, Senhu Wang

## Abstract

Despite ample research on the prevalence of specific psychiatric disorders during COVID-19, we know little about how the pandemic affects the general wellbeing of a wider population. The study investigates the prevalence and predictors of general psychiatric disorders measured by the 12-item General Health Questionnaire (GHQ-12) and frequency of loneliness during COVID-19 in the United Kingdom, a country heavily hit by the pandemic. We analyzed 15,530 respondents of the first large-scale, nationally representative survey of COVID-19 in a developed country, the first wave of Understanding Society COVID-19 Study. Results show that 29.2% of the respondents score 4 or more, the caseness threshold, on the general psychiatric disorder measure, and 35.86% of the respondents sometimes or often feel lonely. Regression analyses show that those who have or had COVID-19-related symptoms are more likely to develop general psychiatric disorders and are lonelier. Women and people in their 20s have higher risks of general psychiatric disorders and loneliness, while having a job and living with a partner are protective factors. This study showcases the general psychiatric disorders and loneliness of broader members of the society during COVID-19 and the underlying social disparities.

## 1. Introduction

As the outbreak of COVID-19 has become a global public health emergency, an increasing number of people are affected psychologically, albeit in various forms and to different degrees (Qiu et al., 2020). Despite ample research on the prevalence and predictors of specific types of psychiatric disorders (Huang and Zhao, 2020; Liu et al., 2020; Voitsidis et al., 2020), little is known about how a national population suffers general psychiatric disorders from the pandemic and feel lonely as a result of such disease control measures as social distancing (Banerjee and Rai, 2020; Galea, Merchant, and Lurie, 2020; Venkatesh et al., 2020), lockdown (Kokou-Kpolou et al., 2020; Tull et al., 2020), and quarantine (Reynolds et al., 2008). General psychiatric disorders and loneliness are arguably more widespread than the specific psychiatric disorders in the past studies (Hwang et al., 2020; Killgore et al., 2020), more likely to be prevalent in the developed countries (Mushtaq et al., 2004), and more severely affecting the socioeconomically disadvantaged groups (Armitage and Nellums, 2020; Berg-Weger and Morley, 2020). Thus, the aim of the study is to explore the prevalence and predictors of general psychiatric disorders and loneliness in the United Kingdom, using the first large-scale, nationally representative survey from a developed country during the pandemic.

The negative impacts of COVID-19, among other infectious diseases, on mental health have been widely documented (Holmes et al., 2020; Duan and Zhu, 2020). Research shows the prevalence of generalized anxiety disorders (Huang and Zhao, 2020), depression (Elbay et al., 2020), insomnia, (Kokou-Kpolou et al., 2020; Voitsidis et al., 2020) and posttraumatic stress symptoms (Liu et al., 2020;) during the COVID-19 and identifies sex (Liu et al., 2020), age (Meng et al., 2020), and neighborhood (Cao et al., 2020) as risk factors. Despite importance of these findings, there is imminent need for a bigger picture of how COVID-19 affects mental health of a wider population more generally.

First, although the widely adopted social distancing, lockdown and quarantine measures may cause severe psychiatric disorders to some people (Galea, Merchant, and Lurie, 2020), much more people bear the emotional burdens of loneliness (Banerjee and Rai, 2020; Hwang et al., 2020; Killgore et al., 2020) and develop minor psychiatric disorders that cannot be fully captured by the specific clinical measures in previous research (Lu et al., 2020). The study alternatively adopts the 12-item General Health Questionnaire that validly predicts a wider range of psychiatric disorders (Goldberg and Williams, 1988) and measures the frequency of loneliness as a signature psychiatric risk posed by the current pandemic (Killgore et al., 2020). It finds much higher prevalence rates, around one-third of the population, on both measures.

Second, while most studies focus on China and developing countries, developed countries are also deeply affected by COVID-19 (Jia et al., 2020). Despite better health care services, the disruptions of social lives are abrupt in the countries where people have always been connected by technologies (Mushtaq et al., 2014). Thus, the residents could have felt lonely and developed the psychiatric disorders in minor yet non-negligible forms. These unique challenges have not been addressed due to the paucity of real-time, large-scale, and nationally representative data. We use the first wave of the Understanding Society COVID-19 Study, a recent, high-quality, nationally representative survey (Institute for Social and Economic Research, 2020) to assess the COVID-19’s impact on 15,530 respondents in the United Kingdom, a developed country heavily hit by the pandemic (Jia et al., 2020).

Third, our knowledge of the psychiatric disorders and loneliness of COVID-19 patients is limited. For example, the percentages of respondents who have been diagnosed with COVID-19 in the previous surveys are lower than the confirmed prevalence rates in those countries (Cao et al., 2020; Huang and Zhao, 2020; Liu et al., 2020), which may result from sampling bias. These rates would also be lower than the predicted prevalence rates after accounting for the difficulty in testing. In some studies, the sample size is too small to draw a meaningful comparison between the COVID-19 patients and the general population (Huang and Zhao, 2020; Liu et al., 2020). In this study, respondents were identified before the pandemic by using the sample of an existing nationally representative survey, and this could help attenuate the selection bias. We also use self-reported COVID-19 symptoms instead of diagnoses to address the problem of delayed testing. Moreover, suspected patients deserve attention in their own rights who suffer stress regardless of confirmation. We find that people who have or had COVID-19 symptoms are more likely to have general psychiatric disorders and are lonelier.

Finally, previous research offers tentative evidence on the impact of COVID-19 on the vulnerable populations such as women and the elderly (Liu et al., 2020; Meng et al., 2020), whose burdens can be assessed with a larger and more representative sample. With a more comprehensive survey, we are able to study other social determinants of psychiatric disorders such as employment status and family structure (Cao et al., 2020; Kawohl and Nordt, 2020). We find that women and people in their 20s are at higher risks of general psychiatric disorders and loneliness, while having a job and living with a partner are protective factors, suggesting social disparities in psychiatric disorders and loneliness.

## 2. Methods

### 2.1. Data

Data for this study come from the first wave of Understanding Society COVID-19 Study, which was conducted from in April 2020 and publicly available on May 29, 2020.

Understanding Society COVID-19 Study a special wave of the UK Household Longitudinal Study (UKHLS), which uses stratified and clustered sampling to provide high-quality and nationally representative panel data of around 40,000 United Kingdom households. The survey procedures were approved by the Ethics Committee of University of Essex. The survey consists of an online questionnaire but those without internet access are interviewed through telephone by trained professionals. A total of 17,450 respondents answered the current survey with an overall response rate of 41.2%. After pair-wise deletion of a small number of missing cases (around 1%), we obtained an analytic sample of 15,530 respondents. To adjust for complex survey design and unequal non-response rates, we use weighting in all analyses (Institute for Social and Economic Research, 2020).

### 2.2. Measures

*Loneliness* is measured by a question adapted from the English Longitudinal Study on Ageing (ELSA). Respondents are asked “In the last 4 weeks, how often did you feel lonely?” The 4-week period is when COVID-19 was widespread in the country. The responses are divided into three categories: “hardly ever or never,” “some of the time,” and “often.”

*General psychiatric disorders* are measured using the 12 items from General Health Questionnaire (GHQ), a validated scale widely used in the community or non-clinical settings (Aalto et al., 2012; Goldberg and Williams, 1988). There are 12 questions about respondents’ depressive, anxiety symptoms, sleeping problems, confidence and overall happiness etc., which were measured on a four-point scale (1 ‘less than usual’, 2 ‘no more than usual’, 3 ‘rather more than usual’, and 4 ‘much more than usual’). Next, 1 and 2 were recoded to 0, and 3 and 4 were recoded to 1. Finally, the values of the 12 questions were then summed, resulting in a scale ranging from 0 (the least mentally distressed) to 12 (the most mentally distressed). GHQ-12 score equal to 4 or more indicates caseness of general psychiatric disorders (Goldberg and Williams, 1988).

*Gender* is a binary variable. *Age* is divided into 5 categories: 18-30, 31-40, 41-50, 51-60, and over 65. Because the United Kingdom consist of four countries, we create a four-category variable for *country of residence:* England, Wales, Scotland and Northern Ireland. Respondents are asked two questions on past and current *COVID-19 symptoms*, “Have you experienced symptoms that could be caused by coronavirus (COVID-19)?” and “Are you currently experiencing symptoms that could be caused by coronavirus?” The answers to the two questions are “Yes” or “No.” *Employment status* and *living with a partner* are also binary variables answered with “Yes” or “No.” For more details about distribution of each variable, see Table 1.

**Table 1.** Sample Characteristics

|  | Column percentages |
| --- | --- |
| Gender |  |
| Male | 41.76 |
| Female | 58.24 |
| Age groups |  |
| 18-30 | 12.47 |
| 31-40 | 13.77 |
| 41-50 | 17.88 |
| 51-64 | 30.17 |
| 65+ | 25.71 |
| Live with a partner |  |
| Yes | 72.47 |
| No | 27.53 |
| Being employed |  |
| Yes | 61.65 |
| No | 38.35 |
| Whether have Covid19 related symptoms |  |
| Never | 87.55 |
| Ever had Covid19 related symptoms | 11.17 |
| Currently Covid19 related symptoms | 1.29 |
| Regions |  |
| England | 80.89 |
| Wales | 5.89 |
| Scotland | 8.91 |
| Northern Ireland | 4.31 |
| N | 15,530 |

Source: Understanding Society COVID-19 Study

### 2.3. Statistical analyses

First, we report the descriptive statistics of the sample. Second, we conduct univariate analyses with T-tests, ANOVA F-tests and Chi-squared tests to explore the differences in general psychiatric disorders and loneliness across different groups. Third, we run multiple regressions to account for covariates. An ordinary-least square regression model was specified for the continuous General Health Questionnaire score, and a logistic regression model was specified to test the odds ratio of falling into the case category. An ordered logistic regression model was specified for the three-category ordinal outcome of loneliness.

## 3. Results

Table 2 shows the prevalence of psychiatric disorders among the whole UK population and sub-population groups, and reports results of T-tests, ANOVA F-tests and Chi-squared tests to compare prevalence of psychiatric disorder across different population groups. First, we find that the population average GHQ-12 score is 2.73 (SD = 3.26) and 29.20% of the population have caseness of psychiatric disorders. However, there are no significant differences between different UK regions in levels of psychiatric disorders. Next, we find that GHQ-12 psychiatric disorder scores and caseness ratio differ significantly across different socio-demographic groups. Specifically, people who ever had or currently have COVID-19-related symptoms tend to have significantly lower GHQ-12 psychiatric disorder scores than those who never had these symptoms. Similarly, 35.81% and 54.50% people who ever had or currently have COVID-19-related symptoms tend to have caseness of psychiatric disorders as opposed to 27.99% of those without COVID-19-related symptoms. In addition, people who are female and younger tend to have significantly higher GHQ-12 psychiatric disorder scores and caseness ratio than their counterparts, suggesting that both groups are particularly at risk of psychiatric disorders. Also, we find that those who live with a partner and employed tend to have significantly lower GHQ-12 psychiatric disorder scores and caseness ratio than those who live alone and unemployed, suggesting that family support and employment may be a potential protective factor of psychiatric disorders.

**Table 2.** Prevalence of General Psychiatric Disorders in the UK during COVID-19

| | GHQ-12 score<br>(M, SD) | T/F tests | GHQ-12<br>caseness<br>ratio (%) | $\chi^2$ tests |
| --- | --- | --- | --- | --- |
| All | 2.73 (3.26) |  | 29.20 |  |
| Regions |  | p = 0.491 |  | p = 0.879 |
| England | 2.73 (3.26) |  | 29.10 |  |
| Wales | 2.75 (3.38) |  | 28.88 |  |
| Scotland | 2.82 (3.31) |  | 30.06 |  |
| Northern Ireland | 2.58 (3.32) |  | 29.70 |  |
| COVID-19-related symptoms |  | p < 0.001 |  | p < 0.001 |
| Never | 2.63 (3.21) |  | 27.99 |  |
| Ever had symptoms | 3.29 (3.46) |  | 35.81 |  |
| Currently have symptoms | 4.89 (4.06) |  | 54.50 |  |
| Gender |  | p < 0.001 |  | p < 0.001 |
| Male | 2.05 (2.89) |  | 21.02 |  |
| Female | 3.22 (3.42) |  | 35.07 |  |
| Age groups |  | p < 0.001 |  | p < 0.001 |
| 18-30 | 3.71 (3.60) |  | 42.36 |  |
| 31-40 | 3.37 (3.59) |  | 37.56 |  |
| 41-50 | 2.86 (3.28) |  | 31.26 |  |
| 51-65 | 2.61 (3.25) |  | 27.34 |  |
| 65+ | 2.00 (2.69) |  | 19.11 |  |
| Live with a partner |  | p < 0.001 |  | p < 0.001 |
| Yes | 2.50 (3.11) |  | 26.31 |  |
| No | 3.35 (3.57) |  | 36.81 |  |
| Employment |  | p = 0.006 |  | p < 0.001 |
| Yes | 2.64 (3.32) |  | 27.17 |  |
| No | 2.79 (3.24) |  | 30.47 |  |
| N | 15,530 |  | 15,530 |  |

Source: Understanding Society COVID-19 Study

Table 3 reports results of Chi-squared tests to explore differences in frequency of loneliness between different socio-demographic groups. Overall, we find that in the last 4 weeks 64.14% of the population report that they have never felted lonely, and 28.63% have sometimes felt lonely and 7.22% have often felt lonely. The frequency of loneliness does not significantly vary across different UK regions. Moreover, we find that people who have experienced or currently have COVID-19 related symptoms, especially the latter group, are significantly more likely feel lonely than those without COVID-19 related symptoms. For example, 20.00% of those with current COVID-19 related symptoms report that they have often felt lonely as opposed to 9.17% of those with previous COVID-19 related symptoms and 6.19% of those without symptoms. In terms of demographic disparities in loneliness, we find that females and younger people tend to have significantly higher frequencies of loneliness than their counterparts, suggesting that both groups are particularly at risk of loneliness during COVID-19. We also find that people who live with a partner and employed tend to have significantly lower frequencies of loneliness than their counterparts, suggesting that family support and employment may prevent people from suffering loneliness.

**Table 3.**
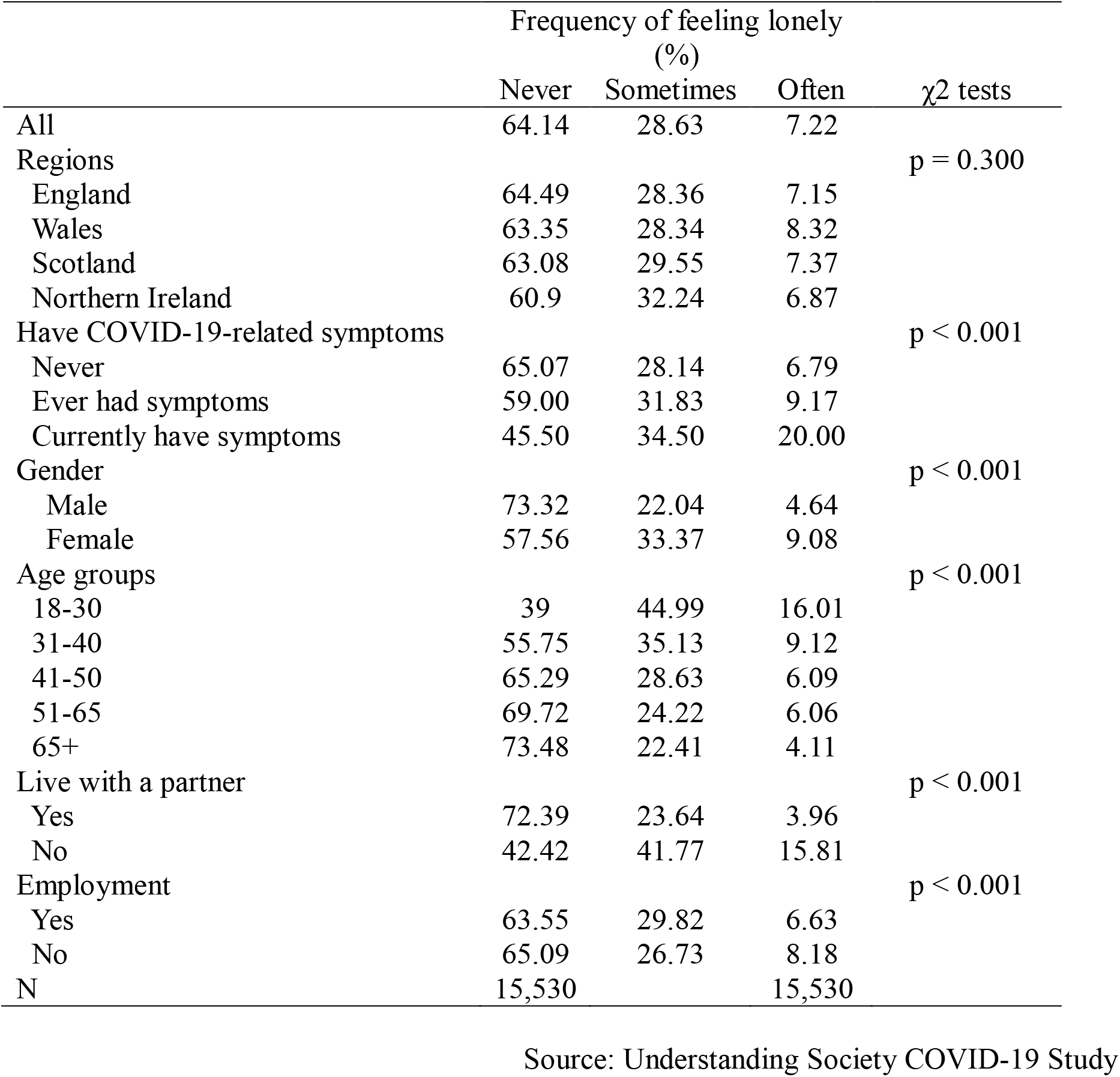
Prevalence of Loneliness in the UK during COVID-19

| | Frequency of feeling lonely<br>(%) | | | $\chi^2$ tests |
| --- | --- | --- | --- | --- |
|  | Never | Sometimes | Often |  |
| All | 64.14 | 28.63 | 7.22 |  |
| Regions |  |  |  | p = 0.300 |
| England | 64.49 | 28.36 | 7.15 |  |
| Wales | 63.35 | 28.34 | 8.32 |  |
| Scotland | 63.08 | 29.55 | 7.37 |  |
| Northern Ireland | 60.9 | 32.24 | 6.87 |  |
| Have COVID-19-related symptoms |  |  |  | p < 0.001 |
| Never | 65.07 | 28.14 | 6.79 |  |
| Ever had symptoms | 59.00 | 31.83 | 9.17 |  |
| Currently have symptoms | 45.50 | 34.50 | 20.00 |  |
| Gender |  |  |  | p < 0.001 |
| Male | 73.32 | 22.04 | 4.64 |  |
| Female | 57.56 | 33.37 | 9.08 |  |
| Age groups |  |  |  | p < 0.001 |
| 18-30 | 39 | 44.99 | 16.01 |  |
| 31-40 | 55.75 | 35.13 | 9.12 |  |
| 41-50 | 65.29 | 28.63 | 6.09 |  |
| 51-65 | 69.72 | 24.22 | 6.06 |  |
| 65+ | 73.48 | 22.41 | 4.11 |  |
| Live with a partner |  |  |  | p < 0.001 |
| Yes | 72.39 | 23.64 | 3.96 |  |
| No | 42.42 | 41.77 | 15.81 |  |
| Employment |  |  |  | p < 0.001 |
| Yes | 63.55 | 29.82 | 6.63 |  |
| No | 65.09 | 26.73 | 8.18 |  |
| N | 15,530 |  | 15,530 |  |

Source: Understanding Society COVID-19 Study

Table 4 uses multivariate regression analyses to explore the net effects of COVID-19 related symptoms and socio-demographic characteristics on psychiatric disorders and loneliness. Model 1 uses OLS model to analyze metrical score of GHQ-12, Model 2 uses logistic regression to analyze GHQ-12 caseness ratio, and Model 3 uses ordered logistic regression to analyze loneliness frequency. Overall, consistent with the findings in Table 2 and 3, the results of multivariate regression models show that there are no significant regional differences in psychiatric disorders and loneliness. Importantly, we find that after controlling for a range of socio-demographic characteristics presence of COVID-19 related symptoms is an important predictor for psychiatric disorders and loneliness with people with current or previous COVID-19 related symptoms being at a greater risk of these mental health problems. Next, with other variables being held constant, females and younger people are still more likely to suffer from psychiatric disorders and loneliness than males and older people, and people who live with a partner and are employed tend to have significantly lower levels of psychiatric disorders and loneliness than those who live alone and are unemployed. These results highlight the importance of COVID-19 related symptoms and continued social disparities in mental health problems during COVID-19 in the UK.

**Table 4.** Predictors of General Psychiatric Disorders and Loneliness in the UK during COVID-19

|  | Model 1<br>GHQ-12<br>score | Model 2<br>GHQ-12<br>caseness ratio | Model 3<br>Loneliness<br>frequency |
| --- | --- | --- | --- |
| Gender (Ref. = Male) | 1.04***<br>(0.08) | 1.86***<br>(0.10) | 1.79***<br>(0.10) |
| Age groups (Ref. = 18-30) |  |  |  |
| 31-40 | 0.06<br>(0.19) | 1.02<br>(0.11) | 0.83<br>(0.09) |
| 41-50 | -0.49**<br>(0.17) | 0.73**<br>(0.07) | 0.57***<br>(0.06) |
| 51-65 | -0.82***<br>(0.15) | 0.59***<br>(0.05) | 0.47***<br>(0.04) |
| 65+ | -1.86***<br>(0.17) | 0.32***<br>(0.03) | 0.29***<br>(0.03) |
| Live a partner (Ref. = Yes) | 0.54***<br>(0.10) | 1.38***<br>(0.09) | 3.22***<br>(0.19) |
| Being employed (Ref. = Yes) | 0.69***<br>(0.13) | 1.36***<br>(0.10) | 1.40***<br>(0.11) |
| Have COVID-19 related symptoms (Ref. = No) |  |  |  |
| Ever had symptoms | 0.63***<br>(0.14) | 1.38***<br>(0.11) | 1.21*<br>(0.10) |
| Currently have symptoms | 1.53***<br>(0.39) | 2.05**<br>(0.46) | 2.10**<br>(0.51) |
| Regions (Ref. = England) |  |  |  |
| Wales | 0.12<br>(0.22) | 1.04<br>(0.14) | 1.29<br>(0.17) |
| Scotland | 0.18<br>(0.14) | 1.08<br>(0.10) | 1.12<br>(0.11) |
| Northern Ireland | -0.15<br>(0.27) | 1.02<br>(0.17) | 1.26<br>(0.22) |
| Constant | 2.46***<br>(0.15) | 0.35***<br>(0.03) |  |
| Constant cut1 |  |  | 2.21***<br>(0.20) |
| Constant cut1 |  |  | 18.23***<br>(1.84) |
| Observations | 15,530 | 15,530 | 15,530 |
| R-squared | 0.08 | 0.05 | 0.10 |

Note. Model 1 used Ordinary Least Square (OLS) regression, Models 2 used logistic regression and 3 used ordered logistic regression. Robust standard errors are in parentheses, \*\*\* p<0.001, \*\* p<0.01, \* p<0.05.
Source: Understanding Society COVID-19 Study

## 4. Discussion

Despite ample research on the prevalence of specific psychiatric disorders during COVID-19, we know little about how the pandemic could affect a wider population in broader ways. The study examines the prevalence and predictors of general psychiatric disorders and loneliness in a developed country. Using nationally representative survey data from 15,530 respondents in the United Kingdom, the study documents the high prevalence rates of general psychiatric disorders (29.2%) and loneliness (35.86%) during the COVID-19 pandemic. It also shows that people with current or past COVID-19 symptoms or various disadvantaged socioeconomic backgrounds are at significantly higher risks of general psychiatric disorders and loneliness. It contributes to the extent literature and informs public health policies in five distinctive ways.

First, the high prevalence rate of general psychiatric disorders lends support to numerous studies on the impact of COVID-19 on mental health (Qiu et al., 2020). It also extends previous research on more specific and severe psychiatric disorders (Huang and Zhao, 2020; Liu et al., 2020; Voitsidis et al., 2020) by showing that nearly one-third of the population are affected by COVID-19 in various forms and to different degrees. Although the minor psychiatric disorders are often less urgent concerns of the public health policies, they are not negligible given the large proportion of the population that have been affected. Only focusing on specific disorders underestimates the psychiatric burdens of the pandemic in more subtle forms and overlooks the needs for psychiatric care of the people who have not been clinically diagnosed.

Second, the prevalence rate of loneliness, an example of minor psychological problem, is high during COVID-19. Over one-third of the respondents sometimes or often feel lonely. This could have arisen from such disease control measures as social distancing, lockdown and quarantine (Tull et al., 2020). It has been repeatedly argued that social isolation during COVID-19 increases loneliness (Killgore et al., 2020), for which the study offers empirical support using a large-scale, nationally representative survey. Loneliness is linked to long-term health outcomes including all-cause mortality (Steptoe et al., 2013), so public health policies need to be aware of the (mental) health consequences of the disease control measures, especially in the developed countries where disruption of social lives could be more abrupt (Jia et al., 2020).

Third, because the respondents were recruited before the pandemic and were asked about COVID-19-related symptoms rather than diagnoses, we obtained a large enough sample size to compare suspected patients with the general population. People with current or past symptoms of COVID-19 are significantly more likely to develop general psychiatric disorders and feel lonelier, evidencing the urgent psychiatric needs of suspected patients.

Fourth, risk factors for psychiatric disorders are examined during the pandemic. Consistent with previous research, being female is a significant predictor of both general psychiatric disorders and loneliness (Liu et al., 2020). Contrary to the popular belief (Armitage and Nellums, 2020; Meng et al., 2020), compared to people in their 20s, older age groups are significantly less likely to feel lonely or develop psychiatric disorders, perhaps because younger people’s economic and social lives are more disrupted by a public health emergency (Cao et al., 2020).

Finally, the study identifies two social determinants of general psychiatric disorders and loneliness during COVID-19. Having a job and living with a partner are both significant protective factors for general psychiatric disorders and loneliness. Moving beyond individual demographics, further studies could explore how social support from work and family buffer the psychological impacts of a pandemic (Cao et al., 2020; Kawohl and Nordt, 2020).

In all, future research and public health policies need to move beyond specific psychiatric disorders to attend to the general psychiatric disorders and loneliness of a larger proportion of the population. They need to pay special attention to vulnerable populations including women, people in their 20s, the unemployed, those not living with a partner, and those who have or had COVID-19 symptoms. A pandemic like COVID-19 could exaggerate social disparities in mental health in subtle ways, calling for research on effective interventions (Holmes et al., 2020; Duan and Zhu, 2020).

## Data Availability

Secondary data can be found at: https://www.understandingsociety.ac.uk/research/themes/covid-19

## Author contributions

The two authors contribute equally.

## Role of the funding source

This research did not receive any specific grant from funding agencies in the public, commercial, or not-for-profit sectors.

## Declaration of Competing Interests

None.

## Notes

### Competing Interest Statement

The authors have declared no competing interest.

### Author Declarations

The original survey was approved by the Ethics Committee of the University of Essex. We analyzed the publicly available, unidentifiable secondary data which does not require IRB approval.

